# Predictive factors of non-pharmacological measures to prevent SARS-CoV-2 contagion: Results of a large adult Spanish sample from the PSY-COVID study

**DOI:** 10.1101/2023.10.30.23297748

**Authors:** Corel Mateo-Canedo, Juan P. Sanabria-Mazo, Jose-Blas Navarro, Igor Cigarroa, María Álvarez, Manuel Armayones, Luis J. Martínez-Blanquet, Rafael Zapata Lamana, Neus Crespo, Marta Korniyenko, Antoni Sanz

## Abstract

**Aim:** the objective of the study was to identify the factors that predicted the intention to carry out coronavirus infection prevention behaviors in the Spanish population during the first wave of the pandemic, a time when the vaccines were in the development phase and the containment of the pandemic depended on non-pharmacological measures. Method: a cross-sectional study (PSY-COVID) was carried out based on an online form that contained instruments for possible predictor variables of seven prevention behaviors, based on a non-probabilistic sampling of 17725 Spanish adults. Results: self-efficacy and outcome expectations related to Coronavirus disease (COVID-19) preventive behaviors, as well as the interaction of perceived severity with both personal vulnerability and actual adherence, appeared as the main predictive variables of the intention to carry out preventive behaviors against the SARS-CoV-2 virus. Slight, but significant differences were observed in behavioral intentions among Spanish autonomous communities, probably linked to differences in the impact of the pandemic in each territory. Conclusion: It is necessary to implement consistent communications that allow for the development of appropriate expectations and encourage adherence to preventive behaviors, as well as recognizing regional disparities in preventive behaviors and their causes to promote compliance.

## INTRODUCTION

Since the first reported case of COVID-19 in China in December 2019, the SARS-CoV-2 virus has spread globally. Spain, one of the earliest and most severely affected countries, experienced significant challenges in implementing public policies to curb the transmission of the virus. As of November 2023, the global number of diagnosed cases exceeded 771 million, with Spain alone recording nearly 14 million cases. The global death toll reached 6.9 million, including 121 thousand fatalities in Spain. The sheer volume of daily infections, hospitalizations, and deaths overwhelmed the healthcare system, posing a persistent challenge throughout the pandemic crisis [1].

In this context, given the absence of pharmacological interventions to prevent contagions before the development of vaccines, several countries implemented health, behavioral, and social measures. In Spain, these measures included home isolation, the mandatory use of face masks, social distancing, and various hygiene protocols at individual, group, and public levels. These actions were taken according to the World Health Organization’s (WHO) recommendations [2], particularly during the first wave of the pandemic. Despite these measures, there has been a substantial disparity in compliance levels between individuals [3], making it necessary to understand the factors contributing to the heterogeneity of adherence to such protective behaviors.

Predictors of compliance with preventive measures have been explored by examining sociodemographic characteristics defining population profiles at risk for non-adherence to public health measures. For instance, evidence suggests that being a woman [4] and older [5] may be associated with higher compliance rates. While some studies have found no direct relationship between compliance and income level [4,6], the findings remain inconclusive. Similarly, the relationship between education level and the willingness to self-isolate requires further investigation [7].

Following prevention and health promotion models [8-11], several studies have analyzed the predictive factors of this type of behavior. For example, Drury et al. [12] highlighted the importance of providing reliable information channels to promote compliance with health recommendations. Beca-Martínez [13] found that adherence to preventive measures was associated with the perceived severity of the virus, trust in specialized information sources, and positive attitudes toward compliance. Additionally, several studies emphasized the role of self-efficacy in complying with measures, along with trust in government and health authorities [14].

Given the unique characteristics of the Spanish population, it was necessary to determine the impact of various factors on compliance with preventive measures. Despite the implementation of nationwide measures, our objective was to analyze the impact of residing in each territorial unit of the country on the adoption of preventive behaviors. Given the exceptional nature of this health alert, the focus was not on a specific set of variables but rather on identifying the predictive capacity among behavioral, cognitive, social, affective, and sociodemographic variables from an exploratory perspective. The assessment tool aimed to capture a broad spectrum of potential predictors of the intention to adhere to health authorities’ recommendations concerning COVID-19.

## METHOD

### Design and procedure

The PSY-COVID study, conducted across 30 countries, aimed to explore the psychological impact of the COVID-19 pandemic. This cross-sectional study gathered data on a broad range of variables from 88740 individuals, and this article reports the findings of a sample of residents in Spain during the lockdown measures. An anonymous online survey (Google Forms^®^) was conducted using non-probabilistic sampling (snowball method). Participants were recruited via various distribution channels, including social networks (Facebook^®^, Instagram^®^, Twitter^®^, WhatsApp^®^), media (newspapers, television, and radio), and institutional contacts (universities, foundations, and health organizations). The survey, administered from May 15th to June 5th, 2020, took approximately 15 minutes to complete. The variables and instruments selected for this survey were validated by a panel of 30 international health researchers and translated into 16 languages.

The survey also included a consent form to participate in this study. This research was approved by the Animal and Human Experimentation Ethics Committee of the Autonomous University of Barcelona (CEEAH-5197) and followed the Code of Ethics of the World Medical Association (Helsinki Declaration) for experiments involving humans.

### Participants

A total of 17,725 individuals responded to the online survey in Spain. However, 2,594 were excluded from this analysis because they resided in other countries during the lockdown measures. Consequently, the final sample for analysis consisted of 15131 participants. The inclusion criteria were: (1) being adults (≥ 18 years old) and (2) being residents in Spain during the lockdown measures implemented during the first wave of the COVID-19 pandemic.

### Measures and instruments

The following sections describe the measures extracted from the PSY-COVID database, which are relevant to this study [15].

### Sociodemographic characteristics

A sociodemographic information questionnaire was included to collect data on age, gender (female and male), educational level (primary, secondary, and university), income level (low, medium, and high), and region of residence in Spain.

### Personality

A brief, ad-hoc version of The NEO Five-Factor Inventory [16] was used to measure the five dimensions of personality: neuroticism, extraversion, agreeableness, conscientiousness, and openness to experience. The highest factor saturation item for each dimension was used. Responses to these items were recorded on a 5-point Likert-type scale, where 1 corresponded to “*strongly disagree*” and 5 to “*strongly agree*.” Higher values indicate greater traits in each dimension.

### Prevention behaviors and cognitions

A 35-item ad-hoc inventory was used to measure seven health behaviors concerning five constructs derived from socio-cognitive models of health prevention (i.e., social cognitive theory [17], planned action theory [18], and the health belief model [19]): (1) post-confinement prevention behavior intention, (2) experience with prevention behaviors, (3) self-efficacy level for prevention behaviors, (4) outcome expectations of prevention behaviors, and (5) social norm of preventive behaviors. The behaviors measured were selected from the prevention recommendations promoted by the WHO and health authorities during the first wave of the pandemic [2]: (1) wearing, carrying, or removing masks; (2) hand washing for at least 40 seconds; (3) avoiding touching the face, mouth, or nose; (4) remembering to wash hands after touching objects; (5) remembering to keep a safe distance from others; (6) resisting the urge to leave home; and (7) asking other people to follow preventive behaviors. Responses to this inventory were recorded on a 4-point Likert-type scale, ranging from 0 (“*not at all*”) to 3 (“*a lot*”). Higher values (ranging from 0 to 21 for each dimension) indicated greater adherence to prevention behaviors. The internal consistency in this sample for post-confinement prevention behavior intention (Cronbach’s *α* = .81), experience with prevention behaviors (Cronbach’s *α* = .72), self-efficacy level for prevention behaviors (Cronbach’s *α* = .78), outcome expectations of prevention behaviors (Cronbach’s *α* = .79), and social norm of preventive behaviors (Cronbach’s *α* = .78) was deemed acceptable to excellent. Additionally, an exploratory factor analysis was carried out for the five scales, revealing a clear unifactorial structure for each, characterized by a single factor with an eigenvalue greater than 1 and an explained variance greater than 40% in all cases.

A 4-item ad-hoc inventory was used to measure adherence to prevention behaviors related to avoiding infection and sanction and proxy control [17] on government, health personnel, and scientists. Responses to this inventory were recorded on a 4-point Likert-type scale, where 1 denoted “*not at all*” and 3 “*a lot*”. Higher values (ranging from 0 to 6 for each dimension) indicated greater engagement in preventive behaviors. The internal consistency in this sample for prevention behaviors to avoid infection (Cronbach’s *α* = .52), prevention behaviors to avoid sanction (Cronbach’s *α* = .72), proxy control of government (Cronbach’s *α* = .59), and proxy control of health personnel and scientists (Cronbach’s *α* = .71) ranged from questionable to acceptable.

A 5-item ad-hoc inventory measured barriers and facilitators for prevention behaviors. The responses to this inventory were recorded on a 5-point Likert-type scale where -2 corresponded to “*makes it very difficult*” and +2 “*makes it very easy*.” Higher values (ranging from -10 to 10) indicated a higher propensity for engaging in preventive behaviors. However, the internal consistency in this sample for barriers and facilitators for prevention behaviors (Cronbach’s *α* = .68) was questionable.

A 4-item ad-hoc inventory was used to measure future threats generated by the pandemic. The responses to this inventory were recorded on a 4-point Likert-type scale where 0 represented “*not at all*” and 3 “*a lot*”. Higher values (ranging from 0 to 12) indicate greater perceived future threats resulting from the pandemic. However, the internal consistency in this sample for future threats (Cronbach’s *α* = .63) was questionable.

A 2-item ad-hoc inventory was used to measure perceived vulnerability to coronavirus for both oneself and others. Responses were recorded on a 5-point Likert-type scale ranging from 0 (“*highly unlikely*”) to 4 (“*highly likely*”). Higher values (ranging from 0 to 4 for each item) indicated greater perceived vulnerability.

A 1-item ad-hoc inventory measured information level (i.e., time spent consulting information on the pandemic). Respondents used a 3-point Likert-type scale where 0 corresponds to “*not at all*,” 2 to “*less than 1 hour*”, 3 to “*1 hour to 2 hours*”, and 3 to “*3 hours or more*”. Higher values (ranging from 0 to 12) indicated more time spent seeking information on the pandemic.

### 4. Data analyses

Statistical analyses were conducted using Stata 17. The descriptive analysis of the sample characteristics was carried out using frequencies (*n*) and percentages (%) for categorical variables and means (*M*) and standard deviations (*SD*) for continuous variables. To maintain the population distribution of each region, data analysis was weighted by the inverse of the ratio of the real population number of a region divided by the number of participants in the sample for that region. Comparison of post-confinement prevention behavior intention means across Spanish regions was conducted through post-hoc comparisons with the type I error corrected using Šidák’s [20] approach. Pearson’s correlation was used to calculate the raw association between measures.

A predictive linear mixed model for post-confinement prevention behavior intention was developed, including the 14 measures of prevention behaviors as independent terms and perceived severity as moderators. A random intercept multilevel model was first estimated using restricted maximum likelihood, with the Spanish region as the random factor. The intra-class correlation (ICC) was also calculated. To assess the significance of the interactions between predictors and the moderator (perceived severity), a chunk likelihood ratio test, as recommended by Kleinbaum et al. [21], was conducted for all interaction terms. In cases where the chunk test was not significant, all interactions were removed. Conversely, if the chunk test was significant, each interaction was individually tested using a Wald test. In instances where a significant interaction was identified, the coefficient of the predictor was estimated for three levels of perceived severity (low = 1000, medium = 9144, and high = 17000).

## RESULTS

### 1. Characteristics of the sample

Sociodemographic information is displayed in Table 1. The sample consisted of 15,131 residents in Spain during the COVID-19 lockdown measures. Of these, 66.8% were female, 62.5% reported medium income levels, and 75% had a university education. The average age was 38.4 years, with the majority residing in the Catalonia region (63.65%).

### 2. Personality and prevention behaviors

Table 1 also provides descriptive statistics for personality traits (i.e., extraversion, conscientiousness, amiability, emotionality, and openness traits), prevention behaviors (i.e., post-confinement behavior intention, past experience behaviors, adherence behavior, reasons for adherence, self-efficacy for behavior, outcome expectations of behavior, barriers/facilitators, and social norms), as well as the level of information, proxy control, perceived vulnerability, and perception of future threats within the sample.

**Table 1.**
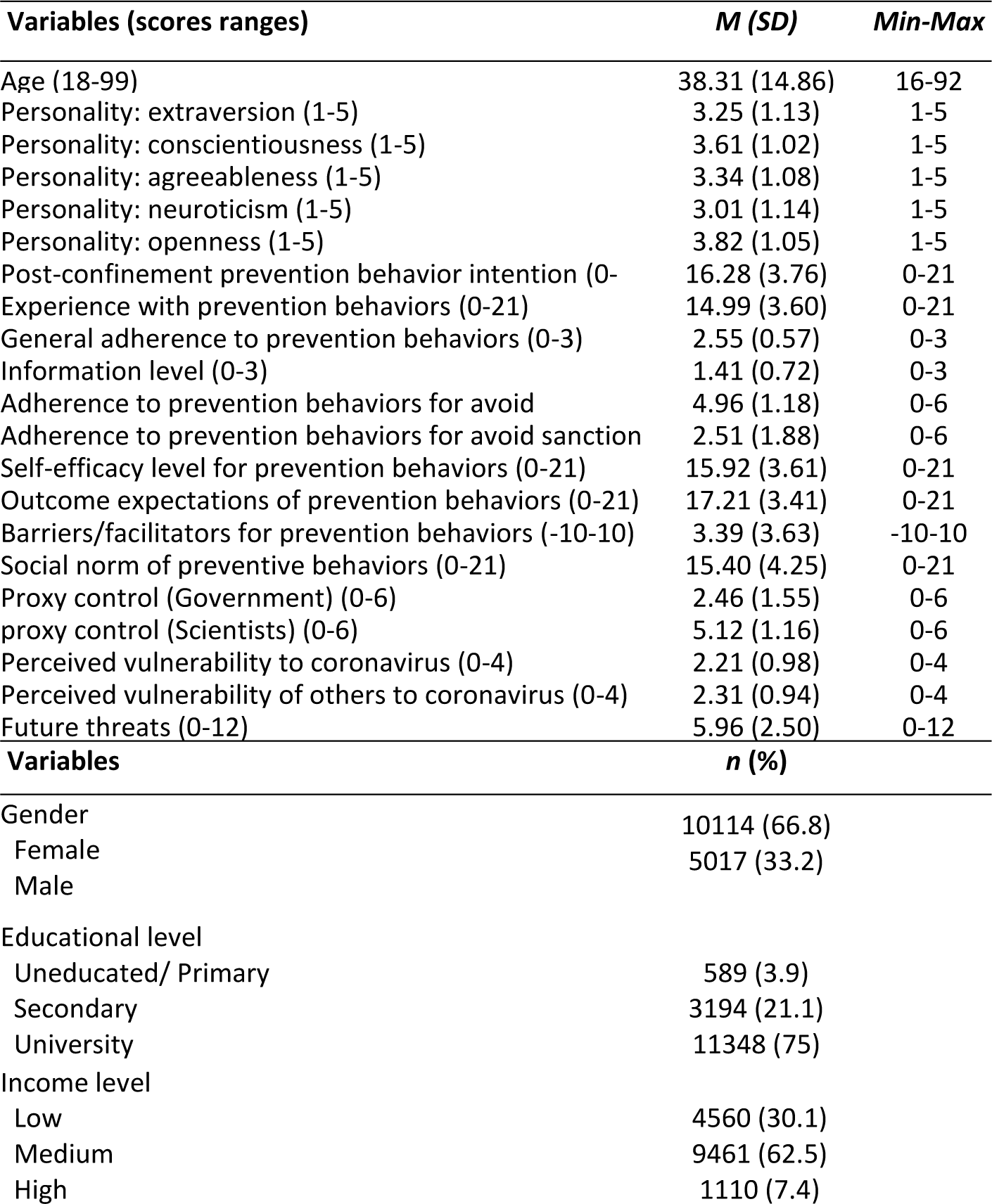
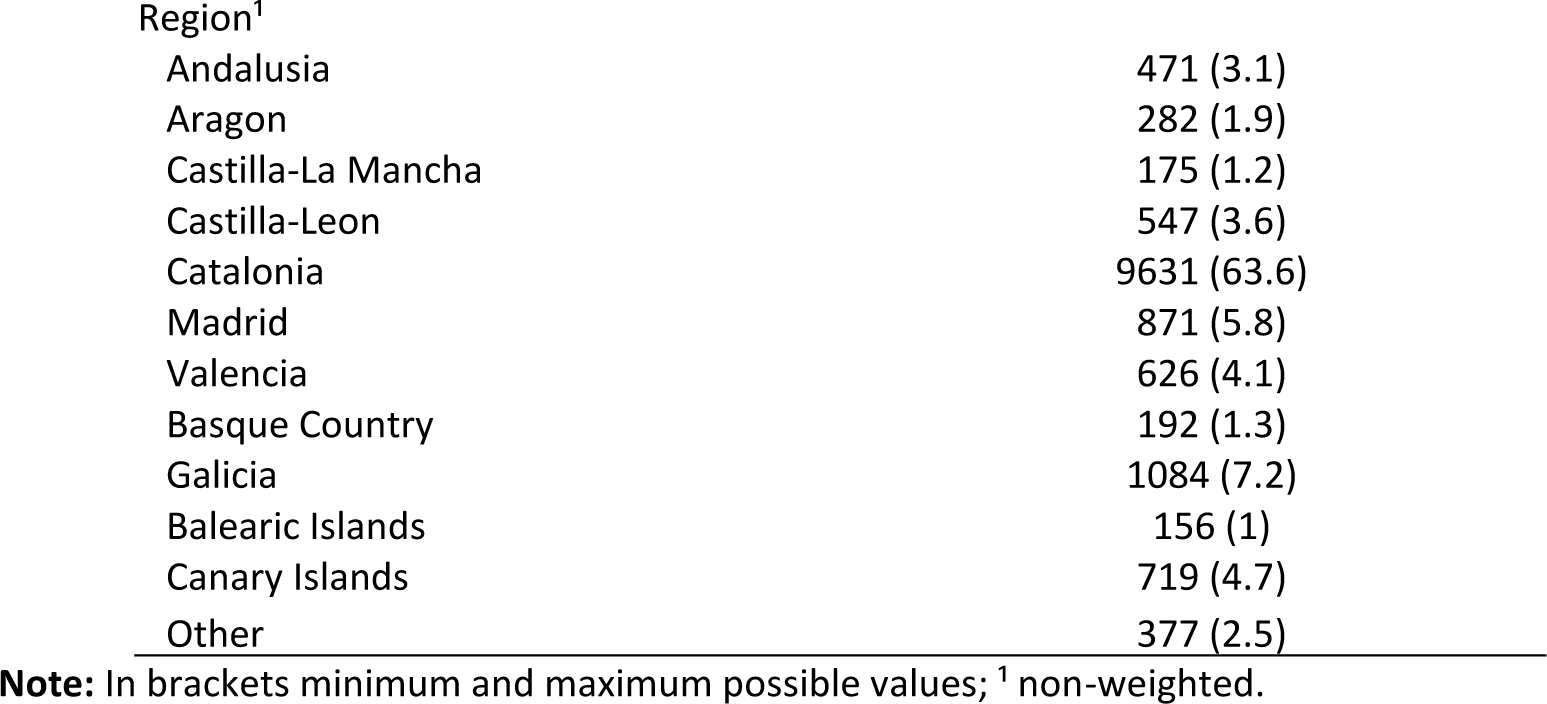
Characteristics of the sample (*N* = 15131)

### 3. Comparison of post-confinement prevention behavior intention between regions

Table 2 shows the means of post-confinement prevention behavior intention separately for each Spanish region. Residents of the Canary Islands showed the highest post-confinement prevention behavior intention compared to those in other regions. In contrast, residents of the Aragón and Balearic Islands region demonstrated the lowest intention for post-confinement prevention behaviors compared to residents of other regions.

**Table 2.**
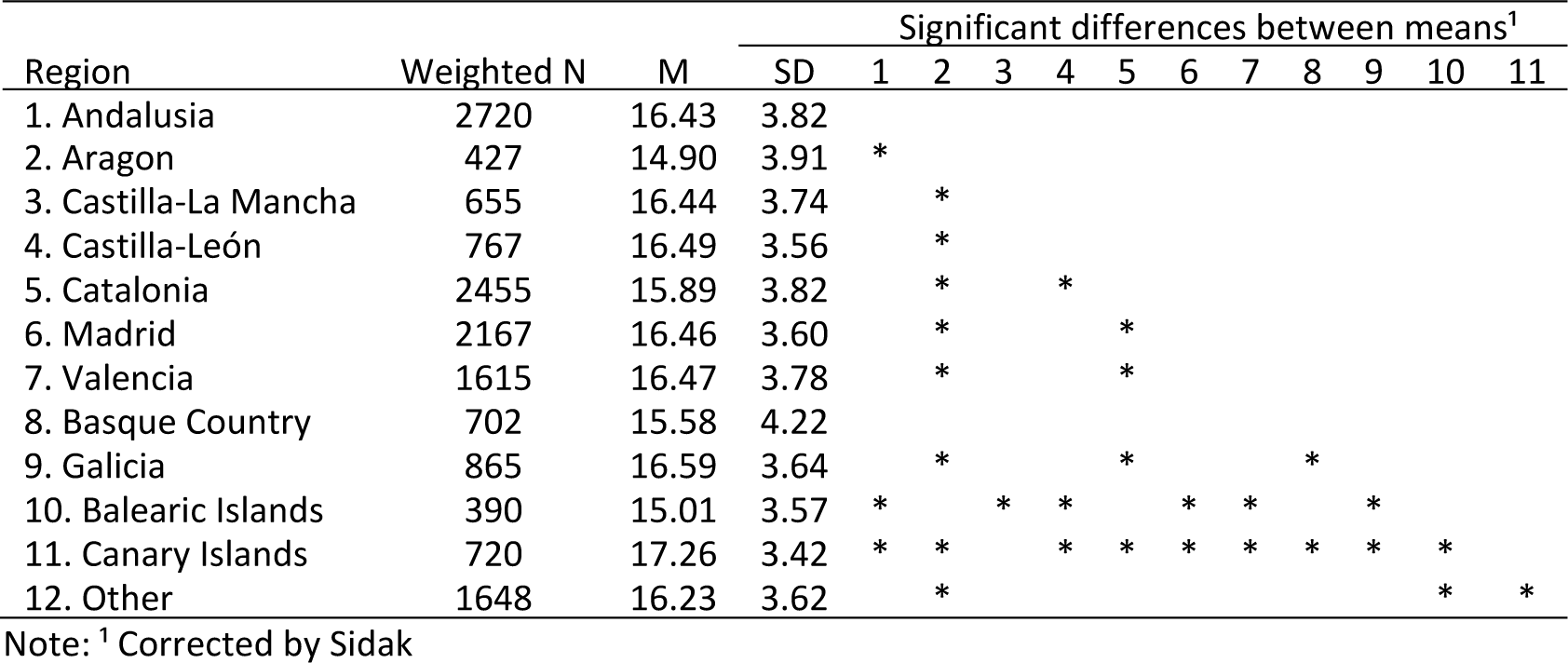
Post-confinement prevention behavior intention comparison between regions.

### 3. Correlations between post-confinement prevention behavior intention and potential predictive factors

Table 3 shows the correlations between post-confinement prevention behavior intention and the sociodemographic factors in the sample of Spanish residents. Among the highest correlation values, the post-confinement prevention behavior intention was notably related to the experience with prevention behaviors (*r* = .64). Both variables were correlated with general adherence (intention *r* = .43; experience *r* = .42), adherence to avoid infection (intention *r* = .51; experience *r* = .52), self-efficacy level (intention *r* = .73; experience *r* = .69), and outcome expectations of prevention behaviors (intention *r* = .72; experience r = .54). Self-efficacy was correlated with outcome expectations (r = .41) and both were related to general adherence (self-efficacy *r* = .42; outcome r = .38) and adherence to avoid infection (self-efficacy *r* = .48; outcome expectations *r* = .49). Finally, social norms correlated with outcome expectations (*r* = .41).

**Table 3.** Bivariate Pearson correlations.

|  | 1 | 2 | 3 | 4 | 5 | 6 | 7 | 8 | 9 | 10 | 11 | 12 | 13 | 14 | 15 | 16 | 17 | 18 | 19 | 20 |
| --- | --- | --- | --- | --- | --- | --- | --- | --- | --- | --- | --- | --- | --- | --- | --- | --- | --- | --- | --- | --- |
| 1 |  |  |  |  |  |  |  |  |  |  |  |  |  |  |  |  |  |  |  |  |
| 2 | -.02 |  |  |  |  |  |  |  |  |  |  |  |  |  |  |  |  |  |  |  |
| 3 | -.00 | .10 |  |  |  |  |  |  |  |  |  |  |  |  |  |  |  |  |  |  |
| 4 | .13 | .32 | .08 |  |  |  |  |  |  |  |  |  |  |  |  |  |  |  |  |  |
| 5 | -.12 | .00 | -.05 | -.07 |  |  |  |  |  |  |  |  |  |  |  |  |  |  |  |  |
| 6 | .09 | .01 | .07 | .09 | .08 |  |  |  |  |  |  |  |  |  |  |  |  |  |  |  |
| 7 | .10 | .04 | .13 | .03 | .08 | .10 |  |  |  |  |  |  |  |  |  |  |  |  |  |  |
| 8 | .08 | .05 | .15 | -.00 | .09 | .12 | .63 |  |  |  |  |  |  |  |  |  |  |  |  |  |
| 9 | .12 | .00 | .12 | .03 | .06 | .05 | .43 | .42 |  |  |  |  |  |  |  |  |  |  |  |  |
| 10 | .23 | .01 | .01 | .05 | .02 | .03 | .20 | .18 | .13 |  |  |  |  |  |  |  |  |  |  |  |
| 11 | .06 | .04 | .12 | -.03 | .07 | .05 | .5 | .52 | .38 | .11 |  |  |  |  |  |  |  |  |  |  |
| 12 | -.11 | .07 | .04 | -.01 | .76 | -.03 | -.05 | -.06 | -.06 | -.03 | .03 |  |  |  |  |  |  |  |  |  |
| 13 | .05 | .00 | .17 | .01 | .03 | .10 | .73 | .69 | .42 | .15 | .48 | -.05 |  |  |  |  |  |  |  |  |
| 14 | .02 | .05 | .11 | .02 | .08 | .07 | .72 | .54 | .39 | .15 | .49 | -.00 | .62 |  |  |  |  |  |  |  |
| 15 | .04 | .12 | .09 | .12 | .02 | .06 | .35 | .25 | .22 | .12 | .29 | .09 | .30 | .40 |  |  |  |  |  |  |
| 16 | .07 | .08 | .09 | .07 | -.05 | .03 | .39 | .26 | .17 | .08 | .24 | .04 | .35 | .41 | .30 |  |  |  |  |  |
| 17 | .03 | .03 | -.00 | .11 | .01 | .08 | .13 | .07 | .12 | .07 | .10 | -.02 | .10 | .16 | .24 | .05 |  |  |  |  |
| 18 | .02 | .08 | .07 | .13 | .01 | .08 | .29 | .21 | .20 | .10 | .24 | -.08 | .24 | .32 | .29 | .17 | .32 |  |  |  |
| 19 | .03 | .05 | -.00 | .02 | .59 | .03 | .17 | .13 | .08 | .07 | .14 | -.07 | .12 | .17 | .08 | .04 | .01 | .10 |  |  |
| 20 | -.03 | .03 | -.00 | -.00 | .07 | .05 | .15 | .12 | .09 | .05 | .12 | -.07 | .12 | .17 | .05 | .00 | .01 | .09 | .80 |  |
| 21 | .01 | -.01 | -.01 | -.03 | .02 | -.01 | -.01 | -.00 | -.01 | .01 | -.02 | .01 | -.01 | -.01 | -.01 | -.03 | -.01 | -.02 | -.01 | -.01 |

Note: 1.Age; 2.Personality: extraversion; 3.Personality: conscientiousness; 4.Personality: amiability; 5.Personality: emotionality; 6.Personality: openness; 7.Post-confinement prevention behavior intention; 8.Experience with prevention behaviors; 9.General adherence to prevention behaviors; 10.Information level; 11.adherence to prevention behaviors for avoid infection; 12.adherence to prevention behaviors for avoid sanction; 13.Self-efficacy level for prevention behaviors; 14.Outcome expectations of prevention behaviors; 15.Barriers/facilitators for prevention behaviors; 16.Social Norm of preventive behaviors; 17. Proxy Control of Government; 18. proxy Control of Scientists-Health staff; 19.Personal Perceived vulnerability to coronavirus (self); 20.Perceived vulnerability to coronavirus (others) ; 21.Future threats.

### 4. Predictive model of the post-confinement prevention behavior intention

Table 4 presents the results of the linear mixed model aimed at predicting post-confinement prevention behavior intention. For a random intercept multilevel model with the Spanish region as a random factor, the ICC was 0.028 (CI 95% 0.012 to 0.061). Despite being a relatively low value, the variance explained by the region (0.049) was statistically significant (*p* = .005), prompting the retention of the random factor in subsequent models. The mean change in coefficient estimates between the model with two sets of adjustment terms and the model with only the four sociodemographic terms was 6.44%, suggesting that the five personality dimensions could be excluded from the model. However, the mean change in coefficient estimates between the model with the sociodemographic set and the model without any adjustment term was 20.25%, highlighting the need to retain the four sociodemographic characteristics in the model.

From the extensive set of variables analyzed, a model composed of the most influential predictors of post-confinement prevention behavior intention was obtained. The variables in this model, ordered by the standardized coefficient *B*, were as follows: self-efficacy (*B* = 0.36; p < .001), outcome expectations (*B* = 0.36; p < .001), and general adherence to prevention behaviors (B = 0.31; p < .01); information level (*B* = 0.25; p < .001), experience with prevention behaviors (*B* = 0.12; p < .001), perceived vulnerability (*B* = 0.12; *p* = .008), proxy control (scientists) (*B* = 0.08; p < .01), social norm of preventive behaviors (*B* = 0.06; *p* < .001), adherence to prevention behaviors to avoid sanction (*B* = -0.03; = .10), barriers/facilitators for prevention behaviors (*B* = 0.02; *p* = .12), future threats (*B* = -0.01; *p* = .23), and proxy control (government) (*B* = 0.00; *p* = .79). Additionally, the perceived severity level adjusted the adherence to prevention behaviors to avoid infection (for low perceived severity, *B* = 0.12; p <= .01; for medium perceived severity, *B* = 0.18; *p* < .001; for high perceived severity, *B* = 0.24; *p* < .001) and perceived vulnerability of others to coronavirus level (for low perceived severity, *B* = -0.80=; *p* = .195; for medium perceived severity, *B* = 0.02; *p* = .60; for high perceived severity, *B* = 0.12; *p* <= .01).

**Table 4.** Mixed model for prediction of post-confinement prevention behavior intention.

| Fixed effects | B | CI 95% (B) | p | $\beta^*$ |
| --- | --- | --- | --- | --- |
| Adherence to prevention behaviors to avoid infection |  |  |  |  |
| For low perceived severity = 1000 | 0.12 | 0.04 to 0.21 | <.01 | 0.04 |
| For medium perceived severity = 9144 | 0.18 | 0.13 to 0.23 | <.001 | 0.06 |
| For high perceived severity = 17000 | 0.24 | 0.20 to 0.28 | <.001 | 0.08 |
| Perceived vulnerability of others to coronavirus |  |  |  |  |
| For low perceived severity = 1000 | -0.08 | -0.20 to 0.04 | .20 | -0.02 |
| For medium perceived severity = 9144 | 0.02 | -0.06 to 0.11 | .60 | 0.01 |
| For high perceived severity = 17000 | 0.12 | 0.05 to 0.19 | <.01 | 0.03 |
| Self-efficacy level for prevention behaviors | 0.36 | 0.33 to 0.40 | <.001 | 0.35 |
| Outcome expectations of prevention behaviors | 0.36 | 0.32 to 0.40 | <.001 | 0.33 |
| General adherence to prevention behaviors | 0.31 | 0.11 to 0.52 | <.01 | 0.05 |
| Information level | 0.25 | 0.15 to 0.34 | <.001 | 0.05 |
| Experience with prevention behaviors | 0.12 | 0.11 to 0.14 | <.001 | 0.12 |
| Perceived vulnerability to coronavirus | 0.12 | 0.03 to 0.20 | <.01 | 0.03 |
| Proxy control (Scientists) | 0.08 | 0.03 to 0.14 | <.01 | 0.03 |
| Social Norm of preventive behaviors | 0.06 | 0.05 to 0.072 | <.001 | 0.07 |
| Adherence to prevention behaviors for avoid sanction | -0.03 | -0.07 to 0.01 | .10 | -0.02 |
| Barriers/facilitators for prevention behaviors | 0.02 | -0.01 to 0.05 | .12 | 0.02 |
| Future threat | -0.01 | -0.02 to 0.01 | .23 | -0.01 |
| Proxy control (Government) | 0.03 | -0.02 to 0.03 | .79 | 0.00 |
| Random effects | B | (B) CI 95% | p |  |
| Spanish region | 0.05 | 0.03 to 0.10 | .01 |  |

Note: Adjusted by age, gender, education level, and income; B\*: standardized regression coefficient

## DISCUSSION

The main purpose of this study was to identify the predictor variables for the intention to comply with COVID-19 preventive and non-pharmacological measures. Additionally, a territorial analysis was conducted to determine the differences in these factors among autonomous communities. The contribution of this study lies in its exploration of how a diverse range of variables impacts the level of compliance with preventive measures in a large sample of Spanish adults. Understanding the psychosocial factors and mechanisms underlying compliance with preventive behaviors to address the transmission of COVID-19 is crucial for designing effective health strategies.

The data collected in this study correspond to the period of the first wave of SARS-CoV-2, which marked a critical juncture in the fight against the COVID-19 pandemic. This period was characterized by the absence of effective pharmacological treatments and vaccines, which were still under development. Amid this context, individuals grappled with significant uncertainty, limited knowledge about the transmission of the virus, and unclear and inconsistent official recommendations for preventive measures [22]. This situation could explain the generally high adherence to reported preventive behaviors, as well as the intention to continue practicing prevention behaviors post-confinement.

The level of proxy control for the government was low, in stark contrast to the trust placed in scientists. The absence of consistent and coherent messaging from the government regarding prevention and containment measures resulted in confusion and diminished levels of trust among the population [22]. Surprisingly, during the initial phase of the pandemic, marked by unprecedented health challenges, a high number of infections and deaths, and the lack of treatments and tools to combat the virus, trust in scientists was significantly high. Roozenbeek et al. [23] found that greater trust in scientists correlated with reduced susceptibility to coronavirus-related misinformation. However, further studies are needed to clarify the dynamics of trust in public authorities.

Among the extensive range of variables analyzed, the model that best predicts the future intention to comply with COVID-19 preventive measures incorporates elements of health promotion models, including self-efficacy, outcome expectations, general adherence to preventive behaviors, previous experience, perceived vulnerability, and social norm. Notably, self-efficacy emerged as the most consistently significant predictor of COVID-19-related behavior. This finding is consistent with several studies identifying self-efficacy as one of the strongest predictors of COVID-19 preventive behaviors [24] and other respiratory syndromes [25]. A systematic review further revealed that self-efficacy was the second most frequent significant predictor (in 87.5% of analyzed studies) of COVID-19-related behavior and intention [26].

Information level was also included in the model, showing a strong relationship with the intention to comply with COVID-19 measures. Shushtari et al. [27] found that investing time in accessing accurate information is pivotal for engaging in effective COVID-19 prevention measures. The internet and social networks are extensively utilized as sources of health-related information [25], having a significant impact on preventive behavior [28]. However, a systematic review [29] revealed that certain sources of information, such as online social networks, contributed to misinformation and eroded trust in the scientific community. Thus, further research will be necessary to identify the specific effects of various forms of information.

Both adherence to prevention behaviors to avoid infection and the perceived vulnerability of others to coronavirus were contingent on the severity level of the pandemic, as measured by Mathieu et al. [30], using data on the number of people infected and the number of deaths caused by the coronavirus. Adherence to preventive behaviors emerged as a predictor of the future intention to engage in these behaviors, with its predictive value increasing as the number of COVID-19 cases and deaths rose. This effect could be attributed to the heightened perception of severity as information about the rise in infections and deaths became more prominent, despite the potential bias in this construct [31]. While few studies have directly examined the impact of epidemiological reports on behavior, several studies suggest that high levels of perceived severity enhance adherence to recommended preventive behaviors [32]. Moreover, it is commonly observed that perceived vulnerability tends to be higher for others than for oneself. For instance, Wilson et al. [33] found that young individuals tended to perceive lower vulnerability for themselves compared to older individuals. Additionally, it has been observed that groups of smokers estimated their likelihood of developing lung cancer only slightly more than groups of non-smokers, with risk assessments being underestimated for both groups [34]. Further research focused on these factors will be essential to comprehensively understand their specific influence.

The Autonomous Community of residence exerted a minor yet significant influence on the obtained results. Following the declaration of the State of Alarm, decisions regarding healthcare measures to combat the virus were centralized at the national level [35]. This centralization is essential for understanding why relatively similar behaviors were observed across diverse regions. Nonetheless, the distribution of infections and deaths showed notable disparities across the Spanish Autonomous Communities. Gutierrez et al. [36] argue that this level of inequality was twice as high as that observed in relation to income distribution, potentially impacting the healthcare system’s capacity during the pandemic. Pre-pandemic records [37] indicated differences in the number of publicly employed healthcare professionals, financed public health expenditures, and hospital beds within the public system, factors that could explain certain disparities in the pandemic’s impact and the preventive behavior of individuals. This study has both limitations and strengths that should be considered when interpreting the results. The extensive and heterogeneous sample, coupled with the wide variety of explored variables, allowed for a comprehensive analysis of the socio-demographic factors influencing individuals’ preventive behavior. However, online data collection excluded a significant percentage of the population without internet access for various reasons, potentially resulting in underrepresentation and diminished statistical power for correlation analyses. Moreover, it is essential to acknowledge the non-probabilistic nature of the sampling method used in this study. However, the impact of this limitation has been largely mitigated by the substantial sample size and the use of weighting in the statistical analyses.

Furthermore, the study’s timing during the pandemic and the government’s stipulated health measures influenced the evaluation of specific factors relevant to that period (such as the utilization of gloves for virus prevention), which later ceased to be the primary focus of analysis. Moreover, certain phenomena that gained significance as the pandemic progressed and had a cumulative effect were not assessed in this study.

Concerning the reliability of the measurement instrument used in this study, it is worth noting that some Cronbach’s alpha values were low. This is because certain measures ad-hoc constructs with a limited number of items, designed to provide a general and exploratory perspective on the variables influencing behavioral intention.

It is also important to note that this study focused specifically on the Spanish population. However, to gain a more comprehensive understanding of the interactions between the severity of the pandemic and the extent of mobility restrictions, it is essential to compare different countries. In this study, differences in preventive behavior were identified, even though the measures in each autonomous community were consistent during the study period. These differences are likely due to varying impacts in each region and structural differences. Conducting comparative analyses across countries will provide insights into how various measures have affected the spread and severity of the pandemic in different regions, aiding in the identification of effective strategies to mitigate its effects. This type of analysis is crucial for informing effective responses to future pandemics and safeguarding public health. The WHO recently emphasized the global establishment of behavioral science units or teams [38], highlighting the urgent need to leverage psychology and behavior design on a global scale, particularly in the context of the COVID-19 pandemic. This initiative positions the creation of these units as central to developing evidence-based policies that not only address immediate societal needs but also enhance preparedness for future health crises.

In summary, this study aimed to identify predictor variables for future compliance with COVID-19 preventive measures among a large sample of Spanish adults during the ‘first wave’ of the pandemic. The findings emphasized the importance of socio-cognitive factors such as self-efficacy and outcome expectations as primary predictors of COVID-19-related behavior, while also recognizing the impact of perceiving the pandemic’s severity on individuals’ intent to adhere to preventive measures. This research offers valuable insights for public health authorities, enabling them to formulate strategies and policies to encourage compliance with preventive measures.

Implications for Public Health Authorities and Media:

- Promoting Self-Efficacy: Unlike hard-to-influence variables such as personality, self-efficacy is a malleable perception or belief that can vary according to various sources of information. Public health authorities should prioritize interventions that enhance individuals’ self-efficacy regarding preventive behaviors, since building confidence in adopting and sustaining these measures is crucial for continued compliance.
- Communication Consistency: Media outlets play a significant role in disseminating information from both government and scientific authorities. Consistent and clear messaging is essential to reduce confusion and build trust among the population, ultimately encouraging adherence to preventive measures.
- Regional Disparities: Addressing regional disparities in preventive behavior and healthcare resources is vital. Tailoring public health strategies to the specific needs and challenges of different autonomous communities can improve overall compliance and contribute to more effective pandemic management.

## Data Availability

All data produced are available online at https://figshare.com/articles/dataset/PSY-COVID-1_Dataset/21695561

## Ethical approval acknowledgment

This research was approved by the Animal and Human Experimentation Ethics Committee of the Autonomous University of Barcelona (CEEAH-5197) and followed the Code of Ethics of the World Medical Association (Helsinki Declaration) for experiments involving humans.

## REFERENCES

1. Verelst F, Kuylen E, Beutels P. Indications for healthcare surge capacity in European countries facing an exponential increase in coronavirus disease (COVID-19) cases, March 2020. Euro Surveill. 2020;25(13). 10.2807/1560-7917.ES.2020.25.13.2000323.

2. Advice for the public on COVID-19 – World Health Organization. Who.int.

3. Siebenhaar KU, Köther AK, Alpers GW. Dealing with the COVID-19 infodemic: Distress by information, information avoidance, and compliance with preventive measures. Front Psychol. 2020;11:567905. 10.3389/fpsyg.2020.567905

4. Uddin S, Imam T, Khushi M, Khan A, Ali M. How did socio-demographic status and personal attributes influence compliance to COVID-19 preventive behaviors during the early outbreak in Japan? Lessons for pandemic management. Pers Individ Dif. 2021;175. 10.1016/j.paid.2021.110692

5. Brouard S, Vasilopoulos P, Becher M. Sociodemographic and psychological correlates of compliance with the COVID-19 public health measures in France. Can J Polit Sci. 2020;53(2):253–8. 10.1017/s0008423920000335

6. Milad E, Bogg T. Spring 2020 COVID-19 surge: prospective relations between demographic factors, personality traits, social cognitions and guideline adherence, mask wearing, and symptoms in a US sample. Annals of Behavioral Medicine. 2021;55(7):665–76. 10.1093/abm/kaab039

7. Plohl N, Musil B. Modeling compliance with COVID-19 prevention guidelines: the critical role of trust in science. Psychol Health Med. 2021;26(1):1–12. 10.1080/13548506.2020.1772988

8. Ajzen I. Understanding attitudes and predicting social behavior. 1980.

9. Bandura A. Self-efficacy: Toward a unifying theory of behavioral change. Psychol Rev. 1977;84(2):191–215. 10.1037/0033-295x.84.2.191

10. Deci EL, Ryan RM, Deci EL, Ryan RM. Conceptualizations of intrinsic motivation and self-determination. Intrinsic motivation and self-determination in human behavior. 1985;11–40.

11. Prochaska JO, DiClemente CC. Stages and processes of self-change of smoking: toward an integrative model of change. J Consult Clin Psychol . 1983;51(3):390–5. 10.1037//0022-006x.51.3.390

12. Drury J, Carter H, Ntontis E, Guven ST. Public behavior in response to the COVID-19 pandemic: understanding the role of group processes. BJPsych Open. 2020;7(1):e11. 10.1192/bjo.2020.139

13. Beca-Martínez MT, Romay-Barja M, Ayala A, Falcon-Romero M, Rodríguez-Blázquez C, Benito A, et al. Trends in COVID-19 vaccine acceptance in Spain, September 2020‒May 2021. Am J Public Health. 2022;112(11):1611–9. 10.2105/AJPH.2022.307039

14. Carlucci L, D’Ambrosio I, Balsamo M. Demographic and attitudinal factors of adherence to quarantine guidelines during COVID-19: The Italian model. Front Psychol . 2020;11:559288. 10.3389/fpsyg.2020.559288

15. Sanz A, Comendador L, Carmona-Cervelló M, Mateo-Canedo C, Sanabria-Mazo JP, Feliu-Soler A et al. PSY-COVID-1 Dataset. 2022. Doi:10.6084/m9.figshare.21695561

16. Manga D, Ramos F, Morán C. The Spanish norms of the NEO Five-Factor Inventory: New data and analyses for its improvement. International Journal of Psychology and Psychological Therapy. 2004;4(3):639–48.

17. Bandura A. The social foundations of thought and action: A social cognitive theory. Englewood Cliffs, NJ: Prentice Hall; 1986.

18. Ajzen I. Attitudes, personality, and behavior. Milton-Keynes: Open University Press/McGraw-Hill; 2005.

19. Carpenter CJ. A meta-analysis of the effectiveness of health belief model variables in predicting behavior. Health Commun . 2010;25(8):661–9. 10.1080/10410236.2010.521906

20. Sidak Z. Rectangular confidence regions for the means of multivariate normal distributions. J Am Stat Assoc . 1967;62(318):626. 10.2307/2283989

21. Kleinbaum DG, Kupper LL, Nizam A, Muller K, Rosenberg ES. Applied Regression Analysis and other Multivariable Methods. 5th ed. Boston (MA): Cengage Learning, Inc; 2014.

22. Royo S. Responding to COVID-19: The case of Spain. Eur Pol Anal . 2020;6(2):180–90. 10.1002/epa2.1099

23. Roozenbeek J, Schneider CR, Dryhurst S, Kerr J, Freeman ALJ, Recchia G, et al. Susceptibility to misinformation about COVID-19 around the world. R Soc Open Sci. 2020;7(10):201199. 10.1098/rsos.201199

24. Karl JA, Fischer R, Druică E, Musso F, Stan A. Testing the effectiveness of the Health Belief Model in predicting preventive behavior during the COVID-19 pandemic: The case of Romania and Italy. Front Psychol . 2021;12:627575. 10.3389/fpsyg.2021.627575

25. Alsulaiman S, Rentner T. The health belief model and preventive measures: A study of the Ministry of health campaign on Coronavirus in Saudi Arabia. J Int Crisis Risk Commun Res. 2018;1(1):27–56. 10.30658/jicrcr.1.1.3

26. Zewdie A, Mose A, Sahle T, Bedewi J, Gashu M, Kebede N, et al. The health belief model’s ability to predict COVID-19 preventive behavior: A systematic review. SAGE Open Med . 2022;10:20503121221113668. 10.1177/20503121221113668

27. Shushtari ZJ, Salimi Y, Ahmadi S, Rajabi-Gilan N, Shirazikhah M, Biglarian A, et al. Social determinants of adherence to COVID-19 preventive guidelines: a comprehensive review. Osong Public Health Res Perspect . 2021;12(6):346–60. 10.24171/j.phrp.2021.0180

28. Amodan BO, Bulage L, Katana E, Ario AR, Fodjo JNS, Colebunders R, et al. Level and determinants of adherence to COVID-19 preventive measures in the first stage of the outbreak in Uganda. Int J Environ Res Public Health . 2020;17(23):8810. 10.3390/ijerph17238810

29. Rocha YM, de Moura GA, Desidério GA, de Oliveira CH, Lourenço FD, de Figueiredo Nicolete LD. The impact of fake news on social media and its influence on health during the COVID-19 pandemic: a systematic review. Z Gesundh Wiss . 2021;31(7):1–10. 10.1007/s10389-021-01658-z

30. Mathieu E, Ritchie H, Rodés-Guirao L, Appel C, Giattino C, Hasell J. Coronavirus pandemic (COVID-19). Our world in data. 2020.

31. Saletti Cuesta L, Tumas N, Berra S. Percepción de riesgo ante el coronavirus en la primera fase de la pandemia en Argentina. Hacia Promoc Salud . 2021;26(1):163–78. 10.17151/hpsal.2021.26.1.13

32. Luo Y, Cheng Y, Sui M. The moderating effects of perceived severity on the generational gap in preventive behaviors during the COVID-19 pandemic in the US. International journal of environmental research and public health. 2011;18(4). 10.3390/ijerph18042011

33. Wilson RF, Sharma AJ, Schluechtermann S, Currie DW, Mangan J, Kaplan B, et al. Factors influencing risk for COVID-19 exposure among young adults aged 18-23 Years-Winnebago County. Morb Mortal Wkly Rep. 2020;2020:1497–502. 10.15585/mmwr.mm6941e2

34. McKenna FP, Warburton DM, Winwood M. Exploring the limits of optimism: the case of smokers’ decision making. Br J Psychol . 1993;84 (Pt 3):389–94. 10.1111/j.2044-8295.1993.tb02490.x

35. BOE-A-2018-10752 Real Decreto-ley 7/2018, de 27 de julio, sobre el acceso universal al Sistema Nacional de Salud. www.boe.es. [cited 2023 Oct 29]. https://www.boe.es/buscar/doc.php?id=BOE-A-2018-10752

36. BOE-A-2020-3692 Real Decreto 463/2020, de 14 de marzo, por el que se declara el estado de alarma para la gestión de la situación de crisis sanitaria ocasionada por el COVID-19. www.boe.es. [cited 2023 Oct 29]. https://www.boe.es/buscar/act.php?id=BOE-A-2020-3692

37. Gutiérrez M-J, Inguanzo B, Orbe S. Distributional impact of COVID-19: regional inequalities in cases and deaths in Spain during the first wave. Appl Econ . 2021;53(31):3636–57. 10.1080/00036846.2021.1884838

38. New global resolution calls for establishment of behavioural science units or teams. www.who.int. [cited 2023 Oct 29]. Available in: https://www.who.int/europe/news/item/20-06-2023-new-global-resolution-calls-for-establishment-of-behavioural-science-units-or-teams.

